# A COVID-19 outbreak in a rheumatology department upon the early days of the pandemic

**DOI:** 10.1101/2020.06.05.20107011

**Authors:** Vasco C. Romão, Filipa Oliveira-Ramos, Ana Rita Cruz-Machado, Patrícia Martins, Sofia Barreira, Joana Silva-Dinis, Luís Mendonça-Galaio, Helena Proença, José Melo Cristino, Ema Sacadura-Leite, Nikita Khmelinskii, José Carlos Romeu, João Eurico Fonseca, on behalf of CHULN Rheumatology Department

## Abstract

**Objectives:** To describe our experience with a coronavirus disease 2019 (COVID-19) outbreak within a large rheumatology department, early in the pandemic.

**Methods:** Symptomatic and asymptomatic healthcare workers (HCWs) had a naso-oropharyngeal swab for detection of severe acute respiratory syndrome coronavirus 2 (SARS-CoV-2) and were followed clinically. Reverse transcription polymerase-chain reaction (RT-PCR) was repeated to document cure, and serological response was assessed. Patients with risk contacts within the department in the 14 days preceding the outbreak were screened for COVID-19 symptoms.

**Results:** 14/34 HCWs (41%; 40±14 years, 71% female) tested positive for SARS-CoV-2, and 11/34 (32%) developed symptoms but were RT-PCR-negative. Half of RT-PCR-positive HCWs did not report fever, cough, or dyspnoea before testing, which were absent in 3/14 cases (21%). Mild disease prevailed (79%), but 3 HCWs had moderate disease requiring further assessment, which excluded severe complications. Nevertheless, symptom duration (28±18 days), viral shedding (31±10 days post-symptom onset, range 15-51) and work absence (29±28 days) were prolonged. 13/14 (93%) of RT-PCR-positive and none of the RT-PCR-negative HCWs had a positive humoral response, with higher IgG-index in individuals over 50 years (14.5±7.7 vs 5.0±4.4, p=0.012). Of 617 rheumatic patients, 8 (1.3%) developed COVID-19 symptoms (1/8 hospitalisation, 8/8 complete recovery), following a consultation/procedure with an asymptomatic (7/8) or mildly-symptomatic (1/8) HCW.

**Conclusions:** A COVID-19 outbreak can occur among HCWs and rheumatic patients, swiftly spreading over the presymptomatic stage. Mild disease without typical symptoms should be recognised, and may evolve with delayed viral shedding, prolonged recovery, and adequate immune response in most individuals.

Key messages
- High infection rates have been reported in healthcare workers (HCWs) dealing with COVID-19 patients. Less is known about potential transmission by pre/asymptomatic carriers, which may be important in the context of rheumatology practice, among both HCWs and patients.
- A COVID-19 outbreak in a rheumatology department affected 41% of HCWs, with fast spreading throughout the presymptomatic stage.
- Mild disease without typical symptoms was common, especially in early phases, evolving with delayed viral shedding (unto 51 days), prolonged recovery (average one month), and adequate immune response (93%) in most individuals.
- Transmission by mostly asymptomatic HCWs occurred to a minority of patients with rheumatic and musculoskeletal diseases (RMDs), who had a globally favourable outcome.
- Asymptomatic and mild disease forms among HCWs should be recognised. Assertive infection control measures and testing strategies are warranted to prevent subclinical contagion between HCWs and patients with RMDs.

## Introduction

Following the initial descriptions in early January 2020 of a novel form of severe pneumonia in patients from Wuhan, China,^1-4^ the coronavirus disease 2019 (COVID-19) quickly spread at a global level. On January 30, the World Health Organization declared it a public health emergency of international concern,^5^ subsequently updated to pandemic on March 11.^6^ After the first reported case in Portugal (March 2), exponential growth led to major restrictive measures.^7^

Consistently high infection rates among healthcare workers (HCWs) have been reported in several hard-hit countries such as China,^8,9^ Italy,^10^ Spain,^11^ or the United States,^12^ despite adequate safety measures.^13^ One possibility is that in-hospital transmission, among patients and HCWs, might be a key form of contagion.^9,14,15^ This is particularly relevant given the transmission dynamics of severe acute respiratory syndrome coronavirus 2 (SARS-CoV-2), whereby presymptomatic/asymptomatic contamination is likely to play a major role in disease spreading.^14-19^

In the early days of the pandemic, most focus was given to severe clinical pictures,^2,8,9,20^ while reports on mild or asymptomatic disease were scarce.^21,22^ This may have contributed to an initial oversight of more general, less severe manifestations, such as upper respiratory and digestive symptoms.^23^ These milder disease forms might be easily undervalued, including by HCWs responding to the pandemic. In healthcare facilities, this may facilitate the generalised spread among HCWs, who can serve as disease-transmission agents.^9,14-16^ Such fact may be particularly relevant in outpatient-oriented departments with a high volume of clinical activity (e.g., rheumatology). In addition, rheumatology practice requires daily close physical contact with patients with rheumatic-musculoskeletal diseases (RMDs), who are often immunosuppressed and have an increased infectious risk.

In the present report, we aim to describe our experience with a COVID-19 outbreak within our department, upon the initial weeks of the pandemic, highlighting clinical, virological and immunological outcomes of HCWs and RMD patients.

## Methods

### Outbreak characterisation

Over the week of 9-15 March, several HCWs of our rheumatology department developed mild symptoms compatible with COVID-19. All staff (symptomatic/asymptomatic) underwent screening for SARS-CoV-2 on 15-16 March. Double naso-oropharyngeal swabs were obtained, and samples were tested for SARS-CoV-2 by reverse transcription-polymerase chain reaction (RT-PCR; cobas®SARS-CoV-2 kit, cobas®6800 System, Roche Diagnostics, USA). All the confirmed and suspected cases were quarantined and referred to public health authorities. Daily remote clinical monitoring of HCWs was conducted by 2 asymptomatic rheumatologists, in conjunction with public health and occupational medicine specialists. Testing of HCWs was repeated (i) 7-14 days after the first negative test in subjects with persisting symptoms; (ii) 5-7 days following the resolution of fever and improvement in respiratory symptoms in confirmed cases.^24^ Two consecutive negative tests were required to confirm viral shedding cessation and allow return to work.^25^ Immunological response to SARS-CoV-2 was evaluated by chemiluminescent immunoassay (MAGLUMI®800 CLIA System, MAGLUMI®2019-nCoV (SARS-CoV-2) IgM/IgG-kits, Snibe Co., Ltd., China) in all HCWs, following symptom resolution and double-negative RT-PCR in confirmed cases.

We contacted patients observed during the previous 2 weeks in the day care unit, outpatient clinic and procedures room, who had possible contacts with confirmed RT-PCR-positive HCWs. Each patient was screened for suggestive symptoms and requested to remain in isolation for 14 days post-contact with the department. Patients with symptoms compatible with COVID-19 were referred to the national health system hotline and signalled to health authorities, who had also received the list of screened patients.

### Study procedures

All HCWs of the rheumatology department who were working during 2-13 March, including visiting fellows, were invited to participate in this study. A standardised questionnaire was administered to collect demographic data, symptom characterisation, disease course and outcome, treatment, comorbidities, and concomitant therapy. Results of laboratory and imaging studies performed, including RT-PCR and IgG/IgM for SARS-CoV-2 were reviewed. Disease course was classified as mild, moderate (requiring physical examination and laboratory/imaging studies) or severe (requiring hospitalisation). Moreover, patients observed in the department between 2-13 March who developed symptoms suggestive of COVID-19 had an appointment scheduled, upon definite resolution, for clinical observation. The same data were collected as for HCWs, in addition to variables related to the RMD and associated treatment. Patients observed in the period of interest who did not develop COVID-19 symptoms, or did so outside the 14-day window after the last contact with the department, were excluded. All study participants signed a study-specific informed consent. This study was approved by the Lisbon Academic Medical Centre Ethics Committee (reference 171/20).

### Statistical analysis

Demographic and clinical characteristics were presented as frequency, mean±standard deviation or median (interquartile range [IQR]), as applicable. Comparison of continuous variables between HCW groups was performed using Kruskal-Wallis (3 groups) or Mann-Whitney U-test (2 groups). Categorical variables were compared using Chi-square or Fisher’s exact test. Agreement between RT-PCR and serological tests was done using Kappa-statistic. Pearson correlation was applied to study the relation of IgG humoral response and clinical variables. Statistical analyses were performed using Stata-12.1 for Mac (StataCorp, College Station, USA) and GraphPad-Prism-7 for MacOS (GraphPad Software, USA). P-value was considered significant at p<0.05.

## Results

### Clinical and virological course of HCWs

A total of 25/34 HCWs (17 rheumatologists, 8 residents, 4 visiting fellows, 1 nurse, 1 health aid, 2 secretaries, 1 cleaning aid) developed symptoms suggestive of a viral infection, 14 of whom had a positive RT-PCR for SARS-CoV-2 (Table 1, Figure 1, Supplementary Figure 1). Ten out of 14 (71%) positive cases were female or younger than 50 years old. Only 4/14 (29%) subjects had a previous history of cardiovascular disease and/or metabolic syndrome, whereas 3/14 (21%) had a diagnosis of immune-mediated inflammatory disease (IMID; one of whom treated with methotrexate 15mg/week). Importantly, 5/14 (36%) HCWs did not develop fever, which lasted ≤3 days in 4/9 (44%) remaining cases. Cough was also absent in the same proportion (36%). Of note, 7/14 (50%) subjects did not develop any of the manifestations of the typical COVID-19 triad prior to the positive RT-PCR test, which were completely missing in 3 cases (21%) throughout the disease. In turn, milder symptoms were already present during the week prior to the outbreak identification in several instances. Anosmia and dysgeusia were present in over half the cases, including 1 subject (HCW6) who did not develop fever, cough or dyspnoea.

**Table 1.**
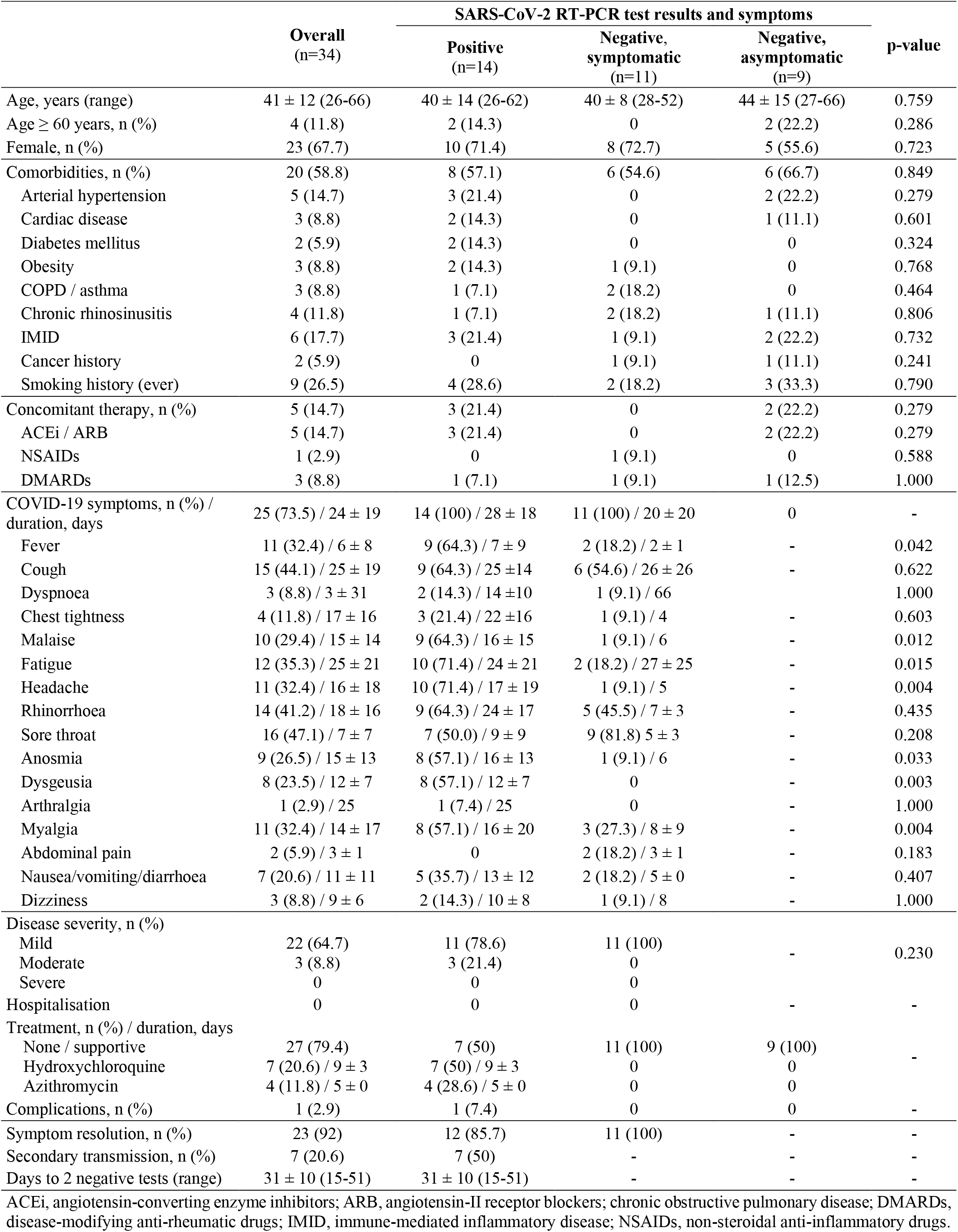
Demographic and clinical characteristics of study participants.

**Figure 1.**
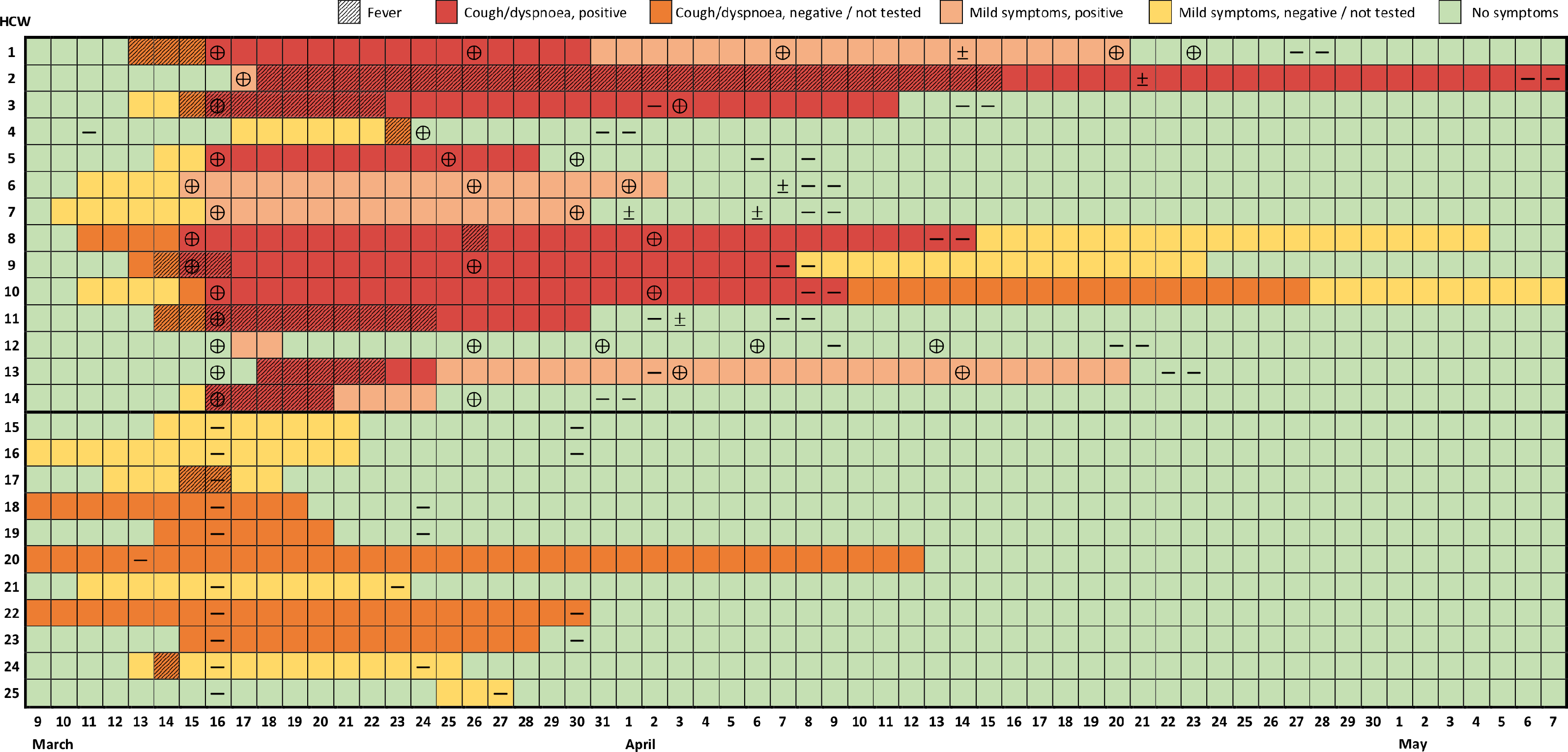
Evolution of symptoms and RT-PCR test results of healthcare workers with confirmed or suspected COVID-19. Each row represents a healthcare worker (HCW) followed over time (columns). Results of reverse transcription polymerase-chain reaction (RT-PCR) are indicated as positive (⊕), negative (—) or indeterminate (±). Mild symptoms include symptoms other than fever, cough and dyspnoea, as referred to in the text and Table 1.

The majority of cases (11/14, 79%) had a benign course. There were no hospitalisations, but 3/14 HCWs (age category 40-70, with relevant comorbidities) underwent clinical, laboratory and radiographic evaluation 7-12 days after symptom onset, due to persistent fever, cough, chest pain and/or shortness of breath. Lymphopaenia (1/3), thrombocytopaenia (1/3), raised lactate dehydrogenase (1/3), D-dimers (2/3), fibrinogen (2/3), and C-reactive protein (CRP; 2/3) were identified, but hypoxaemia and radiographic signs of COVID-19 pneumonia were absent. Seven subjects (50%) were treated with hydroxychloroquine (400mg/day, median 9 days, range 7-14 days) and 4 received concomitant azithromycin (500mg/day, 5 days). One HCW developed a bacterial sinus infection, treated with amoxicillin/clavulanate. Secondary transmission to household members was confirmed in 7/14 (50%) cases, one of which to 2 relatives, all with mild disease.

Despite the favourable course of most cases, symptom duration was prolonged (median 24.5 days, IQR 15-39, range 2-58; Table 1, Figure 1, Supplementary Figure 1). At the end of follow-up, 2 subjects had persistent symptoms (Figure 1, Supplementary Figure 1). Likewise, naso-oropharyngeal RT-PCR remained positive on average for 31±10 days from symptom onset (median 29.5 days, IQR 25-35, range 15-51; Figure 1). This resulted in the need to repeat RT-PCR tests frequently, with a median number of tests per positive subject of 5 (IQR 4-6, range 4-9). Of note, 8/34 (24%) of repetition tests in completely asymptomatic subjects were positive. Yet, this was less than in individuals who repeated testing while still showing some symptoms (13/25, 52%, p=0.024; Figure 1). On average, HCWs were away from work for 29.2±9.8 days (median 24.5, IQR 23-36, range 16-51).

Eleven subjects developed various symptoms but tested negative, even upon retesting (Table 1, Figure 1). These HCWs reported complaints of cough (55%), rhinorrhoea (45%), sore throat (82%), and other symptoms in similar frequency and duration to confirmed cases, over the same time frame. However, fever, fatigue, malaise, headache, myalgia, anosmia, and dysgeusia were significantly less common. Notably, these HCWs had a comparable demographic and comorbidity profile to those with positive RT-PCR and the 9 asymptomatic subjects with negative RT-PCR (Table 1).

No HCWs reported travel from areas with active community transmission. A resident (HCW6), wearing a surgical mask, observed a suggestive case in the emergency department 3 days before symptom onset (March 8), who did not fulfil testing criteria at the time (travel from endemic area). A consultant (HCW4) had a short, unprotected contact on the week preceding the outbreak with an inpatient from another department, later found to have COVID-19. Of note, 7/14 of infected HCWs had a common link to our rheumatological procedures unit, having spent there the most hours over the 2 previous weeks. Nonetheless, the remaining RT-PCR-positive HCWs had minimal exposure to this facility and 2 rheumatologists (HCW18/21), who spent over 10h/week in the unit, tested negative. Finally, all but 10 HCWs (5 RT-PCR-positive, 5 RT-PCR-negative) were present, unprotected, in a 2.5-hour departmental meeting (March 10) addressing the local response to the pandemic. At the time, only 1 HCW (HCW7) had symptoms (mild rhinorrhoea).

### Immunological response

After a median (IQR) of 45 (40.5-48.5) days following symptom onset (or the first RT-PCR test, for asymptomatic subjects), 32 HCWs had an assessment of the serological response (Figure 2). A positive IgM and IgG index (>1.0AU/mL) was seen in, respectively, 2/14 (14.3%) and 13/14 (92.9%) of the confirmed RT-PCR-positive cases and none of the symptomatic/asymptomatic RT-PCR-negative subjects (Figure 2A,B). Both tests had a 96.9% agreement in case classification (Kappa coefficient 0.936). Assessment timing was similar for the HCWs with borderline-positive IgM (HCW11/12, 1.10-1.18AU/mL) or IgG (HCW12/14, 1.10AU/mL) compared to other RT-PCR-positive subjects. In addition, HCW10 had an IgG index below the positive threshold, despite 2 positive RT-PCR tests, no immunosuppression, and comparable evaluation timing and clinical course.

**Figure 2.**
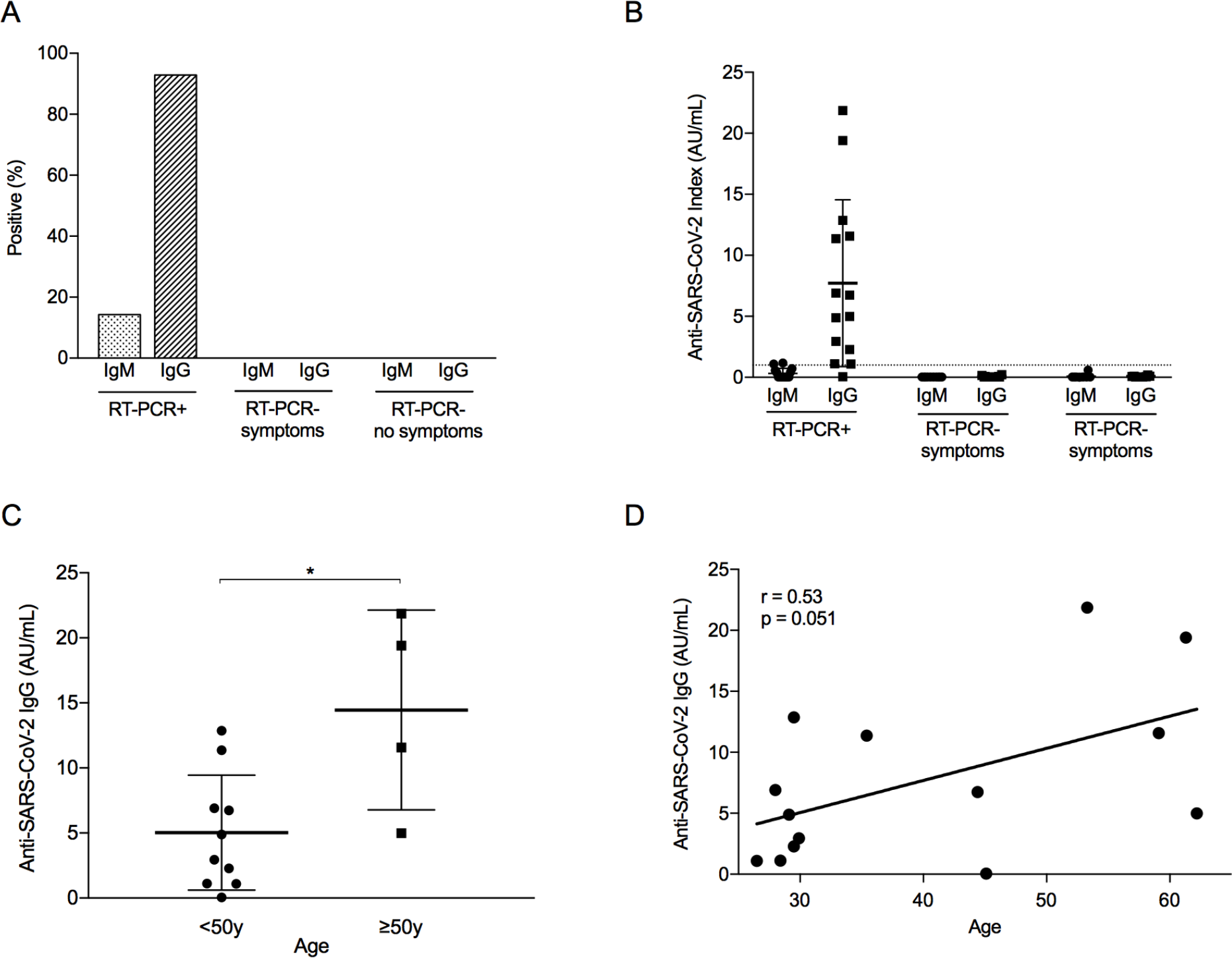
Immunological response to SARS-CoV-2 infection among healthcare workers. **A**. Percentage of healthcare workers (HCW) with a positive immunological response to severe acute respiratory syndrome coronavirus 2 (SARS-CoV-2) infection, defined as an IgM or IgG index equal or above 1.0AU/mL. Analysis differentiated by HCW group, depending on the presence of COVID-19 symptoms and the result of reverse transcription-polymerase chain reaction (RT-PCR). N=32 (RTPCR+, N=14; RT-PCR– symptoms, n=10; RT-PCR– no symptoms, n=8). **B**. Index of anti-SARS-CoV-2 antibodies (IgG and IgM) according to RT-PCR result and COVID-19 symptoms. Dashed line represents positive threshold (1.0AU/mL). Error bars represent mean with standard deviation. **C**. Distribution of anti-SARS-CoV-2 IgG antibodies in RT-PCR+ HCWs, according to age group (below or above 50 years old). *p=0.012. **D**. Correlation between age and anti-SARS-CoV-2 IgG antibodies in RT-PCR+ HCWs. r, Pearson correlation coefficient.

Within the RT-PCR-positive group, considerable variation was seen in the antibody response (Figure 2B). Notably, subjects over 50 years had a higher mean IgG index (14.5±7.7AU/mL)) than younger individuals (5.0±4.4AU/mL, p=0.012; Figure 2C). Although the numbers are small, the 3 older HCWs who had an IgG index above 10AU/mL presented a more severe disease course, with high fever and cough, and 2 of them had raised D-dimers, fibrinogen and CRP. In contrast, the remainder older HCW had a mild course with limited rhinorrhoea and gastrointestinal symptoms, and developed a lower IgG index (4.99AU/mL). Nevertheless, a positive trend was observed in the correlation between age and IgG index (Pearson r=0.53, p=0.051; Figure 2D). No other clinical factor was associated with antibody response, including sex, treatment, or presence/duration of fever, cough or dyspnoea.

### Secondary transmission to patients with RMDs

A total of 617 patients were identified as having had a potential risk contact, 561 (91%) of whom were contacted by telephone and screened for COVID-19 symptoms starting within the 14-day window (Figure 3). We identified 8 (1.3% of total) female patients (mean age 66.8±14.9 years) who developed symptoms compatible with COVID-19 (Table 2). Six patients had a diagnosis of an inflammatory RMD, 3 were treated with conventional synthetic disease-modifying anti-rheumatic drugs (csDMARDs) and glucocorticoids and 2 with biologic DMARDs (bDMARDs). All contacts took place within the same 2 days (9 and 11 March), all but one were with a confirmed infected HCW, and patients denied additional suspicious contacts. Contact tracing for Patient-1 within the department confirmed it to be limited to a symptomatic physician with negative RT-PCR and serology (HCW24). Importantly, in 7/8 cases, the HCW was asymptomatic at the time of contact, and 1 patient (Patient-6) had a consultation with a physician (HCW7) presenting only mild serous rhinorrhea. Of note, 5/8 contacts were in the context of diagnostic (ultrasound) or therapeutic procedures (mesotherapy), which involved prolonged close physician-patient contact.

**Figure 3.**
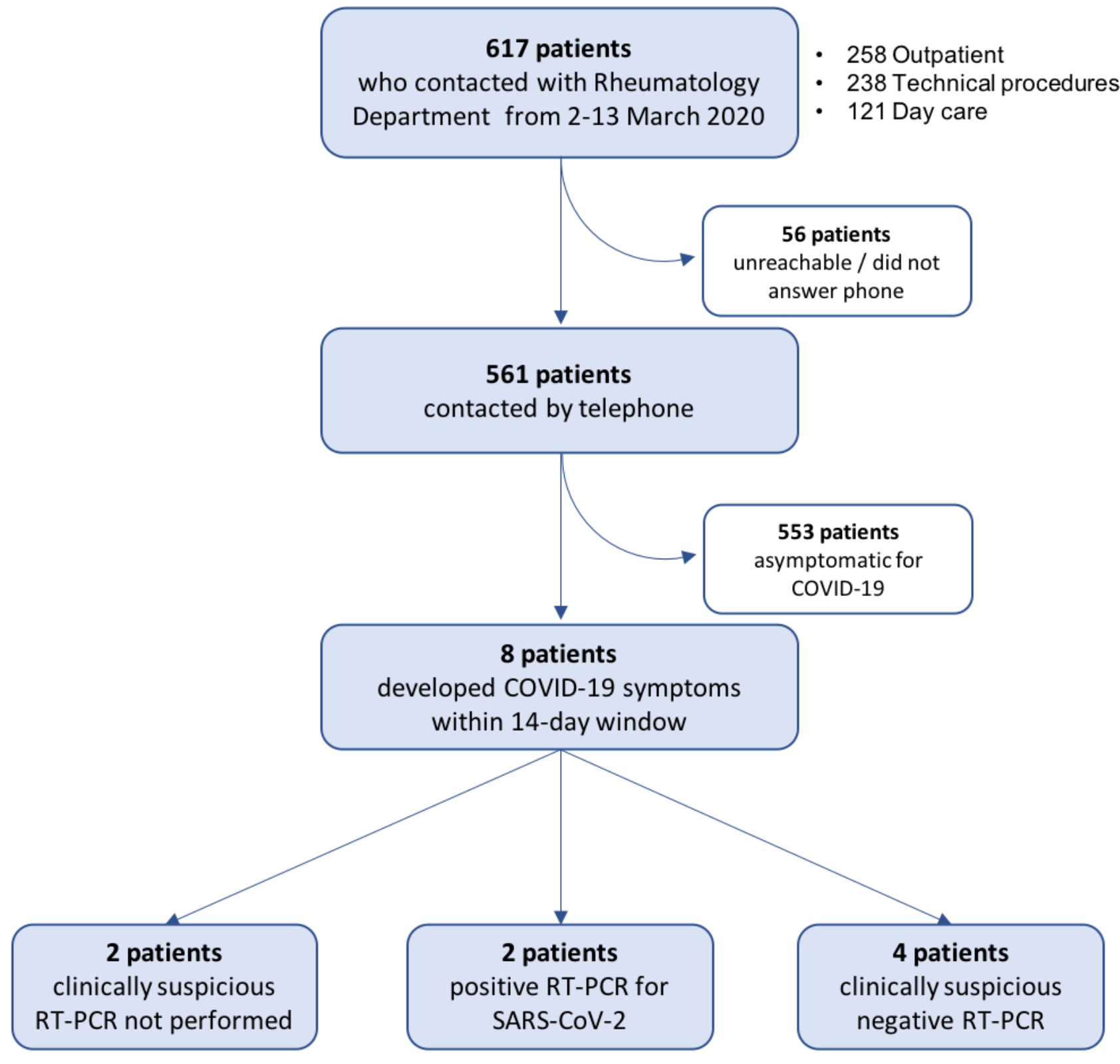
Flow chart of RMD patient screening for symptoms suggestive of COVID-19. Patients with possible contacts with healthcare workers with positive reverse transcription-polymerase chain reaction (RT-PCR) for severe acute respiratory coronavirus 2 (SARS-CoV-2) in the period of 2-13 March 2020 were contacted by telephone and screened for COVID-19 symptoms starting within the subsequent 14 days. See Methods for details.

**Table 2.** Clinical features of RMD patients with confirmed or suspected COVID-19.

| Patient ID | 1 | 2 | 3 | 4 | 5 | 6 | 7 | 8 |
| --- | --- | --- | --- | --- | --- | --- | --- | --- |
| Age category / Sex | 50-60 / F | >90 / F | 60-70 / F | 50-60 / F | 80-90 / F | 70-80 / F | 40-50 / F | 60-70 / F |
| Rheumatic disease | RA/SLE overlap | GCA | Viral reactive arthritis (resolved) | APS | Sarcoidosis | RA | PsA | Rotator cuff tendinopathy |
| Disease duration | 11.1 y | 10.1 y | 7.7 mo | 2.2 y | 13.7 y | 4.7 mo | 15.7 y | 11.2 mo |
| Disease activity | Low | Remission | Remission | No events | Remission | Low | Moderate | Low/mild |
| Comorbidities | HT, heart dx, COPD | HT, CV dx, OP | HT, COPD | HT, CV dx, COPD | HT, heart dx, ILD, CKD | Dyspepsia, spherocytosis | N/A | HT, uterine cancer (past) |
| Smoking status | Past | Past | Past | Active | Never | Never | Never | Never |
| DMARDs (dose, mg) | AZT (150), LEF (20) | MTX (12.5/w), denosumab (60/6mo) | No | No | No | MTX (15/w), HCQ (400) | IFX (3/kg) | No |
| Glucocorticoids (dose, mg) | DFZ (9) | PDN (5) | No | No | No | PDN (7.5) | No | No |
| NSAIDs | No | No | Nimesulide | No | Acemetacin | No | Acemetacin | No |
| ACEi / ARB | Ramipril | No | No | Perindopril | Ramipril | No | No | Lisinopril |
| HCW contact (#) | RT-PCR– (24) | RT-PCR+ (1) | RT-PCR+ (1) | RT-PCR+ (14) | RT-PCR+ (1) | RT-PCR+ (7) | RT-PCR+ (1) | RT-PCR+ (1) |
| Contact date | 9/3/2020 | 11/3/2020 | 9/3/2020 | 9/3/2020 | 9/3/2020 | 11/3/2020 | 11/3/2020 | 11/3/2020 |
| Contact type | US (shoulder) | Mesotherapy (shoulder) | Consultation | Consultation | Mesotherapy (lumbar spine) | Consultation | Mesotherapy (heel) | Mesotherapy (shoulder) |
| Contact duration | 15 min | 10 min | 30 min | 30 min | 10 min | 30 min | 10 min | 10 min |
| Days from contact to symptom onset | 9 | 2 | 4 | 4 | 4 | 3 | 4 | 4 |
| COVID-19 symptoms (order of appearance) | 1) Fv, Mal, Ftg, Hdx, Rhin, Cgh<br>2) Ansm, Dysg | 1) Mal, Ftg, Myalg, Arth, Cgh<br>2) Fv, Hdx, Rhin<br>3) Chx, Dysp, Ansm, Dysg | 1) Mal, Ftg, Hdx, Thr, Cgh, Arth<br>2) Ansm, Dysg<br>3) GI, Dizz | 1) Mal, Hdx, Rhin, Myalg, GI<br>2) Cgh, Ftg<br>3) Ansm, Dysg | 1) Fv, Mal, Fat, Hdx, Rhin, Arth, Myalg, Dizz | 1) Abd, GI<br>2) Rhin, Cgh<br>3) Chx, Hdx | 1) Ftg, Cgh<br>2) Dysg<br>3) Hdx, Abd | 1) Mal, Ftg, Hdx, Thr, Cgh<br>2) Abd, GI<br>3) Fv, Myalg, Dizz |
| Diagnostic RT-PCR test (symptom day) | Positive (13) | Positive (4) | Inconclusive (11)<br>Negative (21) | Negative (9) | N/A | Negative (8) | N/A | Negative (8) |
| Symptom duration | 14 | 20 | 30 | 21 | 31 | 25 | 29 | 28 |
| Hospitalisation (days) | No | Yes (10) | No | No | No | No | No | No |
| ICU admission | N/A | No | N/A | N/A | N/A | N/A | N/A | N/A |
| Complications | No | No | No | No | No | No | No | No |
| Targeted treatment | No | HCQ, LOP+RIT | No | No | No | No | No | No |
| Treatment changes | ↓ DFZ 6mg | Suspended MTX | No | No | ↑ NSAID freq | No | No | No |
| Outcome | Cure | Cure | Cure | Cure | Cure | Cure | Cure | Cure |
| Days to 2 negative tests | 34 | 20 | N/A | N/A | N/A | N/A | N/A | N/A |

Abd, abdominal pain; ACEi, angiotensin-converting enzyme inhibitors; Ansm, anosmia; APS, antiphospholipid syndrome; ARB, angiotensin-II receptor blockers; Arth, arthralgia; AZT, azathioprine; Chx, chest pain/tightness; Cgh, cough; CKD, chronic kidney disease; COPD, chronic obstructive pulmonary disease; COVID-19, coronavirus disease 2019; CV, cerebrovascular; dx, disease; DFZ, deflazacort; Dizz, dizziness/vertigo; DMARDs, disease-modifying anti-rheumatic drugs; Dysg, dysgeusia; Dysp, dyspnoea; Freq, frequency; Ftg, fatigue; Fv, fever; GCA, giant cell arteritis; GI, gastrointestinal, including nausea, vomiting, diarrhoea; HCQ, hydroxychloroquine; HCW, healthcare worker; Hdx, headache; HT, hypertension; ICU, intensive care unit; IFX, infliximab; ILD, interstitial lung disease; LEF, leflunomide; LOP, lopinavir; Mal, malaise; mo, months; MTX, methotrexate; N/A, not applicable; NSAIDs, non-steroidal anti-inflammatory drugs; OP, osteoporosis; PDN, prednisolone; PsA, psoriatic arthritis; RA, rheumatoid arthritis; RIT, ritonavir; Rhin, rhinorrhea; RMD, rheumatic and musculoskeletal diseases; RT-PCR, reverse transcription-polymerase chain reaction; SLE, systemic lupus erythematosus; Thr, sore throat; US, ultrasound; w, weeks; y, years.

Patients developed symptoms on average 4.3±2.1 days (range 2-9) after the contact. Half reported fever, 88% had cough, and only 1 patient reported dyspnoea. General and upper airway symptoms were common, including anosmia (50%) and dysgeusia (63%). Nasopharyngeal swabs were performed in 6/8 cases (8.8±3.1 days post-symptom onset), 2 of which were positive for SARS-CoV-2 and 1 was inconclusive. Two patients were not tested due to difficulty in reaching health authorities or personal choice (self-isolation). Patients with negative/unavailable test still had suggestive COVID-19 symptoms.

All but one patient had a mild-to-moderate course and were clinically recovered after an average of 24.8±5.9 days. A woman in her 90’s with giant cell arteritis and relevant cardiovascular comorbidities, exposed to long-term methotrexate and low-dose glucocorticoids, was hospitalised after 6 days of fever and 3 days of worsening chest pain and dyspnoea. She required oxygen therapy, received a combination of hydroxychloroquine (400mg/day) and lopinavir/ritonavir (800/200mg/day), and was discharged after 10 days. Importantly, none of the patients experienced a flare of the baseline RMD.

## Discussion

Our study provides important lessons on the vulnerability and impact of a COVID-19 outbreak within a large rheumatology department, at a time when universal surgical mask use was not recommended. Over a single week, 41% of HCWs were confirmed to be infected by SARS-CoV-2, and an additional 32% developed mostly overlapping symptoms. While we could not detect the index case, the spread of the contagion was fast and occurred when almost all HCWs were asymptomatic or exhibited only minor symptoms, easily dismissed or attributed to another concurrent viral disease. These findings are in accordance with current concerns around the presymptomatic or asymptomatic transmission of SARS-CoV-2 among HCWs and patients.^9,14-16^ As viral shedding and infectiousness are higher in the 2-3 days prior to symptom onset and rapidly decrease thereafter,^26,27^ a high proportion of contagion occurs during the presymptomatic stage.^18,26^ In addition, asymptomatic^17,22,28,29^ and mild disease forms with limited upper respiratory symptoms are now widely recognised,^23,27^ and may escape vigilance protocols, more focused on the presence of fever, cough, and dyspnoea. This was certainly the case in our cluster, where testing of all HCWs of the department, whether symptomatic or not, was vital to identify cases and contain the outbreak. Therefore, in healthcare settings, continuous mask use, social distancing, and mild-symptom monitoring should be adopted among HCWs, together with proactive testing strategies, to account for potential pre/asymptomatic carriers.^15^

The outbreak had a profound repercussion in the clinical activity of the department. Infected subjects had protracted symptoms and were away from work for around 1 month. In addition, prolonged viral shedding (up to 51 days) led to frequent RT-PCR repetition (median 5 tests) until cure was confirmed, consuming substantial resources. Our findings regarding viral RNA swab positivity are longer than previously reported,^20,30-32^ which may be related to differences in specimen collection (double naso- and oropharyngeal swab in our study), study population, or disease severity. Interestingly, similar nasopharyngeal viral load in patients with mild and severe disease have been reported,^30^ although a separate study concluded otherwise.^33^ Moreover, some data suggest that a positive RT-PCR does not denote the actual presence of viable virus, especially after the first week.^27,34^ However, this is not yet fully established, and we would therefore advocate for 2 consecutive negative tests before HCWs return to work. In effect, 21/59 (36%) of repeat tests were positive, and in 5 instances a positive or indeterminate test followed a first negative result, highlighting the difficulties of interpretation.^34^

All HCWs had a mild-to-moderate disease course and there were no major complications. We believe the positive outcome of the cohort is mainly related to the young mean age, with only 2 HCWs older than 60-years. Alternatively, a lower initial viral exposure load could also explain an overall milder phenotype.^33^ Nevertheless, most of the infected HCWs (93%) developed an immune response, which tended to be more robust in older individuals. This, in turn, may be secondary to a more severe clinical course, known to be strongly associated with age.^20,35^ A possible explanation for this finding could be a higher peak viral load, also previously shown to be positively correlated with age.^30^ Indeed, we highlight that 2/13 (15.4%) IgG-positive HCWs, both under 30-years and with a very mild disease course, had borderline IgG indexes. Nevertheless, other factors, such as T-cell-mediated immunity,^36–38^ may be involved, as 1 HCW in the 40-50-year-old range, who had cough and 2 positive RT-PCR tests, did not develop IgG antibodies 47 days post-symptom onset. Concordance between serology and RT-PCR was otherwise excellent, confirming previous reports.^20,30,31^ Although it cannot be completely excluded, this suggests there were no false-negative RT-PCR results, including in symptomatic HCWs.

Finally, secondary transmission to a minority of patients did occur, from 4 HCWs who were asymptomatic (75%) or had mild upper airway symptoms (25%), mostly in a close proximity contact. As one of the confirmed cases only contacted with HCW24 (negative RT-PCR and serology), we cannot exclude undisclosed community contagion or nosocomial transmission through fomites.^39^ Also, we admit patients with negative/missing RT-PCR could be falsenegative or undiagnosed cases, possibly due to a larger interval between symptom onset and testing. Of note, all contacts occurred when preventive measures had already been adopted and 80% of face-to-face clinical activity had been deferred, which might explain the low number of infected patients. We admit the possibility that contagion could have followed the opposite route (pre/asymptomatic patients-to-HCWs), although symptom timing does suggest otherwise. Notwithstanding the advanced age and long-term use of cs/bDMARDs and glucocorticoids in half the cases, all patients had a favourable outcome. This is in accordance with the largest series to date of patients with IMIDs, failing to show an increase in the incidence of severe disease.^40^

Our study has some limitations. Due to its real-life nature, clinical assessment and RT-PCR timing were clinically-based and differed slightly between subjects. As computed tomography was not performed, we cannot completely exclude COVID-19 pneumonia. Two fellows could not be tested for serology upon finishing their clerkship. Also, 9% of the identified RMD patients could not be reached.

In conclusion, we demonstrate that a COVID-19 outbreak can occur among HCWs and rheumatic patients, spreading over the presymptomatic stage, and evolving with mild-to-moderate symptoms, delayed viral shedding and prolonged recovery.

## Data Availability

Data are available on reasonable request. All data relevant to the study are included in the article or in the supplementary files. Other data are available on reasonable request.

## Acknowledgements

We acknowledge Ms. Sandra Guimarães, Ms. Veneranda Barroca and Ms. Inalda Santos, from the CHULN Rheumatology Department for their assistance to study procedures. We thank Dr Álvaro Ayres Pereira and Dr Tiago Marques, from the CHULN Infectious Diseases Department, for the clinical support in evaluating confirmed and suspicious cases. We thank Dr Clara Almeida, from the CHULN Occupational Health Department, for the contribution regarding serological assay results.

## Contributors

All authors have contributed to study conception and design. VCR, FO-R and JEF designed the project, collected and analysed the data and drafted the manuscript. ARC-M, PM, SB and JS-D contacted patients and obtained the relevant clinical data. LM-G and ES-L coordinated the occupational health response. HP and JMC conducted laboratory testing, including RT-PCR and serology. NK and JCR overviewed clinical monitoring and data collection of healthcare workers. All members of the CHULN Rheumatology Department (collaborative group) actively participated in data acquisition. All authors have critically reviewed the manuscript for important intellectual content, and have read and approved its final version.

## Funding

No specific funding was used for this manuscript.

## Competing interests

All authors declare no competing interests.

## Patient and public involvement

Patients and/or the public were not involved in the design, conduct, reporting or dissemination plans of this research.

## Appendix 1 Collaborative authorship — CHULN Rheumatology Department

Manuel António^1,2^

Pedro Ávila-Ribeiro^1,2^

Rita Barros^1,2^

Raquel Campanilho-Marques^1,2^

Susana Capela^1,2^

Inês Cordeiro^1,2^

Bianca Cristea^1,3^

Eduardo Dourado^1,2^

Luís Gaião^1^

Raquel Freitas^1,4^

Carla Macieira^1^

Joana Martins-Martinho^1,2^

Ana Teresa Melo^1,2^

Carlos Miranda Rosa^1^

Margarida Monteiro^1^

Lila Morena Bueno Silva^1,5^

Lurdes Narciso^1^

Joaquim Polido-Pereira^1,2^

Cristina Ponte^1,2^

Catarina Resende^1^

Maria João Saavedra^1,2^

Fernando Saraiva^1,2^

Rui Lourenço Teixeira^1,2^

Catarina Tenazinha^1,2^

Ana Valido^1,2^

Elsa Vieira-Sousa^1,2^

Pedro Vilas^1,6^

^1^Rheumatology Department, Hospital de Santa Maria, Centro Hospitalar Universitário Lisboa Norte, Lisbon Academic Medical Centre, Lisbon, Portugal.

^2^Rheumatology Research Unit, Instituto de Medicina Molecular João Lobo Antunes, Faculdade de Medicina, Universidade de Lisboa, Lisbon, Portugal.

^3^Internal Medicine 1 Department, Hospital de Santa Maria, Centro Hospitalar Universitário Lisboa Norte, Lisbon Academic Medical Centre, Lisbon, Portugal.

^4^Rheumatology Department, Hospital Garcia de Orta, Almada, Portugal.

^5^Division of Rheumatology, Hospital das Clinicas HCFMUSP, Faculdade de Medicina, Universidade de Sao Paulo, São Paulo, Brazil.

^6^Internal Medicine Department, Hospital de Faro, Centro Hospitalar Universitário do Algarve, Faro, Portugal.

## Notes

### Competing Interest Statement

The authors have declared no competing interest.

### Author Declarations

This study was approved by the Lisbon Academic Medical Centre Ethics Committee (reference 171/20).

## References

1. World Health Organization. Pneumonia of unknown cause - China [Internet]. WHO. 5 January 2020. Available from: https://www.who.int/csr/don/05-january-2020-pneumonia-of-unkown-cause-china/en/

2. Zhu N, Zhang D, Wang W, et al. A novel coronavirus from patients with pneumonia in China, 2019. N Engl J Med 2020;382(8):727–33.

3. Lu H, Stratton CW, Tang YW. Outbreak of pneumonia of unknown etiology in Wuhan, China: The mystery and the miracle. J Med Virol 2020;92(4):401–2.

4. Zhou P, Yang X Lou, Wang XG, et al. A pneumonia outbreak associated with a new coronavirus of probable bat origin. Nature 2020;579(7798):270–3.

5. World Health Organization. Statement on the second meeting of the International Health Regulations (2005) Emergency Committee regarding the outbreak of novel coronavirus (2019-nCoV) [Internet]. WHO. 30 January 2020. Available from: https://www.who.int/news-room/detail/30-01-2020-statement-on-the-second-meeting-of-the-international-health-regulations-(2005)-emergency-committee-regarding-the-outbreak-of-novel-coronavirus-(2019-ncov)

6. World Health Organization. WHO Director-General’s opening remarks at the media briefing on COVID-19 - 11 March 2020 [Internet]. WHO. 11 March 2020. Available from: https://www.who.int/dg/speeches/detail/who-director-general-s-opening-remarks-at-the-media-briefing-on-covid-19-11-march-2020

7. Diário da República. Decreto do Presidente da República 14-A/2020, 2020-03-18 - DRE. 2020;(2):3-5.

8. Wu Z, McGoogan JM. Characteristics of and Important Lessons From the Coronavirus Disease 2019 (COVID-19) Outbreak in China. JAMA 2020;323(13):1239.

9. Wang D, Hu B, Hu C, et al. Clinical Characteristics of 138 Hospitalized Patients with 2019 Novel Coronavirus-Infected Pneumonia in Wuhan, China. JAMA 2020;323(11):1061–9.

10. Remuzzi A, Remuzzi G. COVID-19 and Italy: what next? Lancet 2020;395(10231): 1225-8.

11. Instituto de Salud Carlos III. Informe sobre la situatión de COVID-19 en España. Informe COVID-19 n^o^ 23. 16 de abril de 2020. 2020.

12. Burrer SL, de Perio MA, Hughes MM, et al. Characteristics of Health Care Personnel with COVID-19 — United States, February 12-April 9, 2020. MMWR Morb Mortal Wkly Rep 2020;69(15):477–81.

13. European Center for Disease Prevention and Control. Coronavirus disease 2019 (COVID-19) in the EU/EEA and the UK-ninth update, 23 April 2020. 2020. Available from: https://www.ecdc.europa.eu/sites/default/files/documents/covid-19-rapid-risk-assessment-coronavirus-disease-2019-ninth-update-23-april-2020.pdf

14. Chang D, Xu H, Rebaza A, Sharma L, Dela Cruz CS. Protecting health-care workers from subclinical coronavirus infection. Lancet Respir Med 2020;8(3):e13.

15. Black JRM, Bailey C, Przewrocka J, Dijkstra KK, Swanton C. COVID-19: the case for health-care worker screening to prevent hospital transmission. Lancet 2020;395(20):1418–20.

16. Arons MM, Hatfield KM, Reddy SC, et al. Presymptomatic SARS-CoV-2 Infections and Transmission in a Skilled Nursing Facility. N Engl J Med 2020; NEJMoa2008457.

17. Bai Y, Yao L, Wei T, et al. Presumed Asymptomatic Carrier Transmission of COVID-19. JAMA 2020;323(14): 1406.

18. Li R, Pei S, Chen B, et al. Substantial undocumented infection facilitates the rapid dissemination of novel coronavirus (SARS-CoV2). Science 2020;3221(January):eabb3221.

19. Chan JFW, Yuan S, Kok KH, et al. A familial cluster of pneumonia associated with the 2019 novel coronavirus indicating person-to-person transmission: a study of a family cluster. Lancet 2020;395(10223):514–23.

20. Zhou F, Yu T, Du R, et al. Clinical course and risk factors for mortality of adult inpatients with COVID-19 in Wuhan, China: a retrospective cohort study. Lancet 2020;395(10229):1054–62.

21. Arashiro T, Furukawa K, Nakamura A. COVID-19 in 2 Persons with Mild Upper Respiratory Symptoms on a Cruise Ship, Japan. Emerg Infect Dis 2020;26(6):1345–8.

22. Hoehl S, Rabenau H, Berger A, et al. Evidence of SARS-CoV-2 infection in returning travelers from Wuhan, China. N Engl J Med 2020;382(13): 1278-80.

23. Gandhi RT, Lynch JB, del Rio C. Mild or Moderate Covid-19. N Engl J Med 2020;NEJMcp2009249.

24. Centers for Disease Control and Prevention. Criteria for Return to Work for Healthcare Personnel with Suspected or Confirmed COVID-19 (Interim Guidance), April 30 2020. Available from: https://www.cdc.gov/coronavirus/2019-ncov/hcp/return-to-work.html

25. European Centre for Disease Prevention and Control. Guidance for discharge and ending isolation in the context of widespread community transmission of COVID-19, 8 April 2020. 2020;(April). Available from: https://www.ecdc.europa.eu/en/publications-data/covid-19-guidance-discharge-and-ending-isolation

26. He X, Lau EHY, Wu P, et al. Temporal dynamics in viral shedding and transmissibility of COVID-19. Nat Med 2020;26(5):672–5.

27. Wölfel R, Corman VM, Guggemos W, et al. Virological assessment of hospitalized patients with COVID-2019. Nature 2020. http://doi.org/10.1038/s41586-020-2196-x

28. Rothe C, Schunk M, Sothmann P, et al. Transmission of 2019-NCOV infection from an asymptomatic contact in Germany. N Engl J Med 2020;382(10):970–1.

29. Mizumoto K, Kagaya K, Zarebski A, Chowell G. Estimating the asymptomatic proportion of coronavirus disease 2019 (COVID-19) cases on board the Diamond Princess cruise ship, Yokohama, Japan, 2020. Eurosurveillance 2020;25(10):1–5.

30. To KKW, Tsang OTY, Leung WS, et al. Temporal profiles of viral load in posterior oropharyngeal saliva samples and serum antibody responses during infection by SARS-CoV-2: an observational cohort study. Lancet Infect Dis 2020;20(5):565–74.

31. Wajnberg A, Mansour M, Leven E, et al. Humoral immune response and prolonged PCR positivity in a cohort of 1343 SARS-CoV 2 patients in the New York City region. medRxiv 2020;2020.04.30.20085613.

32. Vinet L, Zhedanov A. Profile of RT-PCR for SARS-CoV-2: a preliminary study from 56 COVID-19 patients. J Chem Inf Model 2010;53(9):287.

33. Liu Y, Yan LM, Wan L, et al. Viral dynamics in mild and severe cases of COVID-19. Lancet Infect Dis 2020;S1473-3099(20):30232-2.

34. Sethuraman N, Jeremiah SS, Ryo A. Interpreting Diagnostic Tests for SARS-CoV-2. JAMA 2020. http://doi.org/10.1001/jama.2020.8259.

35. Yang J, Zheng Y, Gou X, et al. Prevalence of comorbidities and its effects in patients infected with SARS-CoV-2: a systematic review and meta-analysis. Int J Infect Dis 2020;94:91-5.

36. Grifoni A, Weiskopf D, Ramirez SI, et al. Targets of T cell responses to SARS-CoV-2 coronavirus in humans with COVID-19 disease and unexposed individuals. Cell 2020. http://doi.org/10.1016/j.cell.2020.05.015.

37. Braun J, Loyal L, Frentsch M, et al. Presence of SARS-CoV-2 reactive T cells in COVID-19 patients and healthy donors. medRxiv 2020;2020.04.17.20061440.

38. Soresina A, Moratto D, Chiarini M, et al. Two X-linked agammaglobulinemia patients develop pneumonia as COVID-19 manifestation but recover. Pediatr Allergy Immunol 2020. http://doi.org/10.1111/pai.13263.

39. Ong SWX, Tan YK, Chia PY, et al. Air, Surface Environmental, and Personal Protective Equipment Contamination by Severe Acute Respiratory Syndrome Coronavirus 2 (SARS-CoV-2) from a Symptomatic Patient. JAMA 2020;323(16):1610.

40. Haberman R, Axelrad J, Chen A, et al. Covid-19 in Immune-Mediated Inflammatory Diseases — Case Series from New York. N Engl J Med 2020;NEJMc2009567.

